# Point Of Care Thyroglobulin Lateral Flow Immunoassay for Rapid Detection of Differentiated Thyroid Carcinoma Metastasis to Cervical Lymph Nodes

**DOI:** 10.1101/2025.03.12.25323841

**Authors:** Sagi Angel, Uri Yoel, Kathelina Kristollari, Merav Fraenkel, Emily Bosin, Miri Elezra, Tim Axelrod, Robert S. Marks

## Abstract

Metastatic cervical lymph nodes (LN) are detected in 20-30% of patients with differentiated thyroid cancer (DTC). Current guidelines recommend that once a cervical LN is suspected to be DTC metastasis during a neck ultrasound (US) procedure, it should be investigated via a fine needle aspiration (FNA) biopsy for cytological evaluation and saline washout of the needle for thyroglobulin (Tg) measurement (FNA-Tg). Since Tg is a protein produced exclusively by thyroid follicular cells, a positive FNA-Tg result establishes the diagnosis of metastatic DTC irrespective of cytology. The conventional, immunoassay-based, FNA-Tg washout requires a laboratory and skilled personnel. We developed a semi-quantitative, lateral flow-based method which was shown to detect at the point-of-care (POC), within 10 minutes, positive Tg samples in needle washouts of a suspicious LN at the site of FNA biopsy. In the pre-clinical phase, the POC-Tg limit of detection was determined to be at a concentration equal to 5 ng/mL, after a 1 mL dilution with normal saline. Our prototype was optimized by evaluating different components: types of membranes, pads, antibodies, and gold conjugates. We evaluated our POC-Tg kits on thirty clinical samples: 16 were found positive while the other 14 were seen as negative. All the negative and positive results were further validated by the attending clinical labs, resulting in 100% compatibility compared to the standard procedure. The proof-of-value of our POC-Tg test lies in its ability to significantly reduce the time to results, thus enhancing clinical decision-making, and saving time and valuable resources.

## 1. Introduction

The thyroid gland is the largest endocrine gland in the body [1]. It is a highly vascularized organ with a rich lymphatic network located anteriorly in the neck [2]. It consists of two lobes and one isthmus that binds them together [2, 3]. Thyroid hormones, secreted by the thyroid gland, play a critical role in the regulation of multiple metabolic functions, such as cellular metabolism and energy expenditure. In addition, they modulate the function of many organs, such as the heart (e.g., increasing heart rate), the brain (e.g., influencing neural activity) [4, 5].

Differentiated thyroid carcinoma (DTC), which originates from follicular cells of the thyroid, has demonstrated a rising incidence in the last three decades, with a prevalence of approximately 1% in the general population. DTC main histological types include both follicular thyroid carcinoma (FTC) and papillary thyroid carcinoma (PTC). The latter accounts for approximately 85% of all DTCs [6-8]. DTCs typically demonstrate a favorable prognosis after total or near-total thyroidectomy, often accompanied by iodine 131 (I-131) treatment when indicated [9]. Preoperative ultrasound identifies cervical lymph nodes (LNs), suspected of harboring metastases in 20–31% of individuals diagnosed with DTC (particularly PTC). PTC is known to recur in more than 25% of patients, mainly involving cervical LNs, as seen during a long-term follow-up [11]. When a clinically significant metastatic cervical LN is diagnosed, it is typically recommended to consider compartmental neck dissection. This may reduce the risk of recurrence and possibly mortality [11-13].

The main tumor biomarker of DTC is the glycoprotein thyroglobulin (Tg) - a 660 kDa, dimeric glycoprotein produced exclusively by the follicular cells of the thyroid gland [14]. Current guidelines recommend that a suspicious cervical LN, in the context of known or suspected DTC, should be investigated with ultrasound-guided fine needle aspiration (FNA) for both cytological evaluation (FNAC) and Tg measurement (FNA-Tg), using a saline washout of the needle content [12]. Since Tg is a protein produced exclusively by thyroid follicular cells, FNA-Tg was shown to have high specificity (94%) and sensitivity (91%) in the early detection of metastatic PTC to cervical LNs [15], sometimes exceeding the accuracy of FNAC [16-19]. Following the dilution of the FNA-Tg needle content with 1 mL of 0.9% saline [19], a Tg concentration above 10 ng/mL is highly suspicious for LN metastasis, while a concentration below 1 ng/mL indicates a low likelihood of metastatic LN of DTC origin. A Tg concentration between 1-10 ng/mL should be interpreted individually as moderately suspicious. Following the evaluation of an indeterminate or suspicious cervical LN, using US-guided FNAC and FNA-Tg, the sample is sent to the cytology lab and the washout fluid is evaluated using convention enzyme-linked immunosorbent assay (ELISA)-based immunoassay [20]. Beyond the time needed to accomplish the cytological evaluation, the accuracy rate of FNAC taken from cervical LN is 70-85% in most studies [21]. This challenge can be addressed with an accurate point-of-care assay for Tg (POC-Tg) which could rapidly confirm or rule out a diagnosis of metastatic DTC within minutes.

Lateral flow immunoassay (LFIA) is a well-established platform in the field of point-of-care diagnostics [22]. The platform provides real-time results for the detection and semi-quantification of analytes of complex physiological fluids (urine, blood, saliva, etc.). As a self-contained and one-step operation test, it usually requires only a few drops of sample. Results are displayed within the timeframe of 5–30 min [23]. The basic working principle of the LFIA is the capture of the analyte by colloidal gold or colored nanoparticles via Tg-target specific antibodies, creating a “sandwich complex” which exhibits a visible line on a membrane, indicating a positive answer. Commonly, a LFIA strip consists of several key components such as a nitrocellulose membrane, and various pads, all laminated on a backing card and placed in a cassette [24, 25].

Different commercial LFIAs have been available for the detection of tumor protein biomarkers. Nevertheless, cancer biomarker tests are almost exclusively carried out in centralized laboratories, thus restricting the point of care (POC) diagnostics [22, 26]. To address this gap, we have developed and optimized a POC-Tg LFIA that provides rapid “yes or no” results for LNs suspected as DTC metastases, thus enabling to accurately identify metastatic LNs, facilitating quicker treatment decisions. Herein we present the technological and clinical results of our LFIA-based POC-Tg.

## 2. Methodology

### 2.1. Reagents and materials

Reagents: Gold (III) chloride trihydrate (AuCl_3_‘3H_2_O, cat. no. 334049), Trizma base (cat. no. T1503-100G), Polyethylene glycol 15-20K (PEG 15-20K, cat. no. P2263-500G), Sodium azide (NaN_3_, cat. no. BCBT5374), sodium phosphate dibasic (Na_2_HPO_4_, cat. no. 1.06586), Sodium phosphate monobasic dihydrate (NaH_2_PO_4_*2H_2_O, cat. no. 1.06342), Ethylenediaminetetraacetic acid (EDTA, cat. no. 431788-25G), Normal human serum (cat. no. S1-100ML), Polyethylene glycol 8K (PEG 8K, cat. no. P5413-500G) were sourced from Sigma-Aldrich (Merck). Hydrochloric Acid 32% v/v (HCl, cat. no. H/1100/PB15), Sodium chloride (NaCl, cat. no. BP358-1), Ethanol 99% (cat. no. 0600DF/15) were purchased from Fisher. Trisodium citrate dihydrate (Na_3_C_6_H_5_O_7_ · 2H_2_O, cat. no. 1.06448.0500) and Potassium carbonate (K_2_CO_3_, cat. no. 1.04928.0500) were purchased from Emsure. Lactose (cat. no. 17814) was supplied by Fluka, Trehalose dihydrate (cat. no. 28719.290) from VWR, while Tween 20 (Polysorbate 20, cat. no. 233362500) was purchased from Acros. Normal saline solution (0.9% w/v Sodium chloride, 10 mL) was acquired from B. Braun whereas, plastic cassettes (4mm*60 mm) were purchased from Orgenics LTD.

Membranes and pads: Nitrocellulose membranes (cat. no. CN 120; SN12; FF120) were purchased from Unisart; Mdi, and Whatman, respectively. Whatman supplied the absorbent pad (GB003 Gel Blotting paper); the plastic backing card (LP-25) was obtained from Mdi and the conjugate filter pads (8980 and 6614) were purchased from Ahlstrom Munksjo.

Immunoreagents: Mouse anti-Tg monoclonal antibodies (1.4mg/ml, cat. no. HM392), Anti-Tg mouse monoclonal antibodies (0.89mg/ml, cat. no. HM393), Anti-Tg mouse monoclonal antibodies (1.4mg/ml, cat. no. HM394), Anti-Tg mouse monoclonal antibodies (0.5mg/ml, HM395) were purchased from East Coast Bio. Rabbit IgG (12.8mg/ml, cat. no. AGRIG-0100) and Goat anti-rabbit antibodies (8.76mg/ml, cat. no. ABGAR-0500) were supplied by AristaBio.

### 2.2. Synthesis of AuNP and conjugation to antibodies

#### 2.2.1. Preparation of gold nanoparticles

The gold nanoparticles (AuNPs) were synthesized using trisodium citrate as a reducing agent. Upon boiling 750mL of DDW for 10 minutes in a clean 2L vessel, DDW was poured out, and 570mL of fresh DDW was added and brought to a simmer while stirring. Once it simmered, 4.9mL of the 1% (w/v) AuCl3 was added, followed by dispensing drops of 8.3ml of 1% (w/v) trisodium citrate on it. The solution was allowed to react for 15 minutes, then it was subjected to spectral analysis using a spectrometer to confirm the AuNPs size, concentration, and purity. Absorbance is measured around 400-700nm for 20 nm (520-523 wavelength) diameter AuNPs and around (524-527) for 40 nm AuNPs. Once characterized, the colloidal gold was filtered through a 0.2 µm filter into a sealed container and left overnight to settle. Lastly, the solution was reevaluated via spectral analysis to confirm the lack of possible aggregation.

#### 2.2.2. Conjugation of AuNPs to antibodies

Before conjugating the AuNPs to the antibodies, a cut-off screening evaluation was conducted to establish an optimal concentration-pH ratio. After adjusting the 25 mL AuNPs of 0.1 OD to pH 7 using 0.2M K2CO3, 20 mL of the pH-adjusted solution was transferred to a 50 mL tube. The antibody concentration established by the cut-off evaluation test was brought to a final volume of 1 mL, then added to the tittered gold while swirling at RT for 2 hours, wrapped in aluminum foil to avoid light exposure. Following this procedure, 0.2 mL of 10% (w/v) PEG (15-20K/BS) was diluted with 800 µL DDW and added to the conjugation mixture, while maintaining constant swirling. The tubes were rolled on a carousel for another 50 minutes. Then, 800 µL of 10% (w/v) PEG (15-20k), was added to the solution while mixing. The tubes rolled for another 10 minutes. Freshly prepared conjugates were transferred into Oak Ridge 30 mL tubes and balanced with 0.9 mL of 10 mM gold resuspension buffer at pH 7.5 and centrifugated at 15°C for 45 min at 9,500 rpm in a Sorvall RC5 equipped with an SS34 rotor. The supernatant was discarded using a vacuum system with an attached glass Pasteur pipette. The pellet was resuspended with 1 mL of gold resuspension buffer (GRB) and transferred to a fresh microtube. The GRB was prepared by combining 125 mL of 0.2M Phosphate (NaP) buffer pH=7.4 [25 mM], 2.5 mL of 10% BSA [1%], 2.5 mL of 10% Sodium azide [0.1%] and DDW to a final volume of 25 mL. The 0.2M NaP buffer used to prepare the GRB solution was made by combining 40.5 mL of 0.2M Na_2_HPO_4_ solution and 9.5 mL of 0.2M NaH_2_PO_4_ solution to a final volume of 50 mL and was stored at 4°C. Lastly, 20 µL of the conjugate was diluted in 980 µL DDW and measured using a spectrophotometer at 540 nm for a stock concentration with an OD of approximately 12.

### 2.3. Test development and optimization

Various membranes and conjugation pads were tested for the research and development of POC-Tg. A singular component was tested at a time, and the component that performed best would be selected for the next stage of testing. The process of elimination in finding the optimal conditions for the device was conducted in the following steps:

#### 2.3.1 Membrane testing

Three nitrocellulose membranes (CN; SN12; FF120) were tested for supporting the antibodies flow movement. The SN12 nitrocellulose membrane was from mdi technologies was chosen as the most suitable membrane. After the solutions were dispensed, the membranes were dried in an oven at 50°C for 10 minutes. Membranes were stored at room temperature in a closed box with desiccants until use. Various types of membranes were tested.

#### 2.3.2. Antibody selection and conjugate pad preparation

The conjugate filter pads (8980 and 6614) were tested for storage and release of the antibody-conjugated gold nanoparticles. In this aspect. the 8980 conjugate filter pad from Ahlstrom-Munksjo performed best. The chosen antibody for conjugation in the final version was HM-395. Pads were cut to sizes (25×1cm) and placed over parafilm tape. A conjugation pad solution with AuNP in DDW with a predetermined final concentration was prepared. The conjugate solution-loaded pads were incubated at 37°C for 3 hours. Then they were exposed to 50°C for 1 hour. The conjugate pad solution x2 was prepared using 2.04 g lactose, 5mL of 40% (w/v) sucrose [4%], 10 mL of 10% (w/v) BSA [2%], 1mL of 10% (v/v) Triton x100 [0.2%], 0.5mL of 10% (w/v) sodium azide solution [0.1%], 10ml of 0.5M Tris pH 7.5 [0.1M], 10mL of 10% (w/v) Peg 8000 [2%] and brought to 50ml using DDW. The conjugate pad solution was mixed in equal amounts of AuNPs to form the final solution, x1 conjugate pad solution with the desired OD of the conjugate solution.

#### 2.3.3. Test and control line preparation

The final composition of the dispensed antibodies on the nitrocellulose membrane consisted of an equal mix of clones HM-392, HM-393, and HM-394 using their original concentrations, which brought them to a 1.23mg/ml mix stock solution, which was then diluted to 1mg/ml in the dispensing solution. The antibodies dispensing solution consists of 2% of ethanol 99%; 4% of 50% (w/v) trehalose solution, and 10% of 500mM Tris buffer solution (pH=7.5). According to the concentration of the received antibodies, a final dispensing concentration is made to 1mg/ml by diluting with DDW. Both the test (anti-Tg) and the control lines (anti-antibody) were dispensed from a 1mg/ml final solution concentration via a lateral flow reagent dispenser. The test line was dispensed 10 mm from the near end of the membrane, while the control line was dispensed 18 mm from the near end. The line thickness was 1mm. Rabbit IgG was used for the dispensed control line, while goat anti-rabbit IgG was used for the control line conjugation protocol.

#### 2.3.4. Device composition and assembly

The nitrocellulose (NC) membrane (2.5cm wide) containing immobilized antibodies, was fitted and centered in the appropriate part of a plastic backing card. The conjugate pad was placed at the lower part of the card, with a 2mm overlap between the membranes. The sample pad measuring 2cm in length placed on top of the remaining lower part of the backing card, created an 8mm overlap between itself and the conjugate pad. There was no overlap between the sample pad and the NC membrane. The absorbent pad was placed at the top of the membrane with a 2mm overlap. Upon desiccating overnight, the card is cut into strips of 4mm width and the 4×60 mm strips are then placed in a plastic cassette.

The wash reagent buffer was composed of 1ml 10% (w/v) BSA solution; 100µL 500 mM EDTA; 100µL; 10% (v/v) Triton x100; 100µL 10% Tween 20, 200µL 500mM TRIS (pH=7.5) 100mM, and DDW were mixed to a final volume of 10 ml.

#### 2.3.5. LOD and strength line assessment

To assess the LOD, relations between Tg concentrations, and strength of the signal, the test line has been evaluated and scored according to Figure 2. Aliquots of known concentrations of Tg in PBS were used to evaluate the various combinations of components in the assay. The lines were evaluated and scored from 0: line is not shown to 2: exhibiting a very strong and dark line. A score of 0.1 is used to indicate a weak, but visible positive result. A score under 0.1 (0.05) is considered negative, even if noticeable.

**Figure 1.**
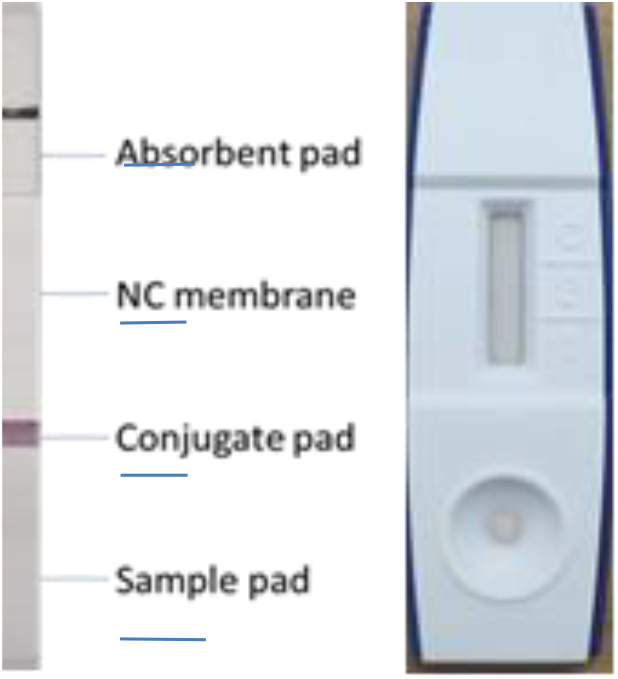
Arrangement of the device’s components listed as absorbent pad, nitrocellulose membrane, conjugate pad, and sample pad. On the device on the right, the round sample well allows the dispensed fluid to flow directly to the sample pad while the rectangular window enables the user to see the test results.

**Figure 2.**
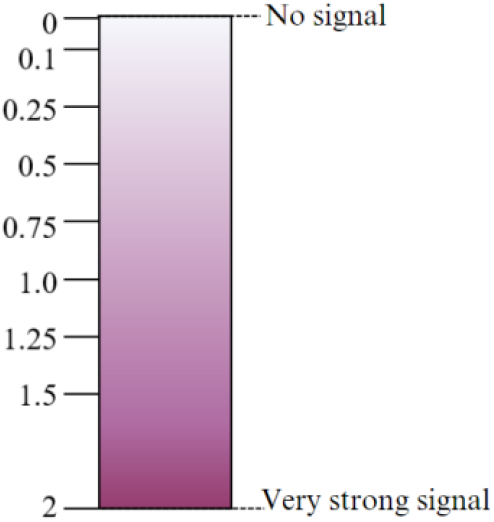
Test line score color legend. Based on this color scheme gradient, the samples were assessed and scored from 0 (no signal) to 2 (very strong signal) depending on the intensity of the signal.

The initial evaluation of the LOD of Tg concentration, as part of assembling the POC-Tg, was done by spiked samples of human serum with known concentrations of Tg. To increase the sensitivity and specificity of the POC-Tg, we used 100 µl 0.9% Saline to dilute samples. Using this dilutional factor, we had 10 times more concentrated samples, when compared with the accepted dilution (for clinical samples) with 1 mL 0.9% Saline. Following the LOD evaluation, we examined the correlation between Tg levels and test line strength in clinical samples to gather additional data on the relationship between these two factors, particularly within the lower ranges of Tg concentrations. This test was done by comparing positive samples taken from the thyroid and several dilutions of those samples prepared using saline 0.9%, to the clinical laboratory gold standard test. Additionally, this test was done in conjunction with the analysis of raw data obtained from collected samples without dilutions. In total, 16 dilutions of samples taken from ten patients were tested.

### 2.4. Data collection and statistical evaluation

Patient demographic and clinical data were not recorded for the present study. However, they are accessible for further analyses at SUMC records. The clinical laboratory validation, as well as the POC-Tg results were recorded and compared. To assess the connection between the strength of the test line score and the concentration of Tg, the samples divided into groups of test line strength and the average corresponding Tg concentration was measured with a 95% confidence interval.

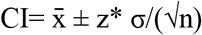

where 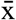 is the sample mean, σ is the standard deviation, n is the sample size, and z* represents the appropriate z*(1.96 for 95% CI).

In order to test significant difference between groups, a Student’s t-test was conducted as follows:

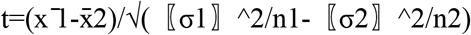

Where 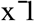 is the mean of sample 1. σ1 is the standard deviation of sample1. n1 is the sample size of sample1.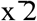 is the mean of sample 2. σ2 is the standard deviation of sample 2. n2 is the sample size in sample 2.

### 2.5. Clinical validation and ethical considerations

#### 2.5.1. Ethical approval

Ethical approval for this study was obtained by the Helsinki committee of Soroka University Medical Center (SUMC), approval number 190-17-SOR. All patients provided written informed consent before the FNA procedure was initiated. Patients 18-year-old and older, able to understand and sign the informed consent form, who were evaluated for cervical LN suspected as DTC metastases, were offered to participate. Pregnant women were excluded.

#### 2.5.2. POC-Tg validation

The evaluation of the POC-Tg kit was conducted concurrently with the routine evaluation of suspected cervical LN (Figure 3.B). The needle content of the passage which was dedicated to Tg-washout was diluted with 100µL 0.9% w/w saline solution (hence, the concentration was 10 times greater than the formal accepted dilution), which was drawn through the needle by which the procedure was done. This amount of 100µL mildly bloody material was then transferred into an Eppendorf tube, from which 40µL were assessed by the POC-Tg for the presence of Tg in the washout fluids. The remaining 60µL was further diluted to a final volume of 600µL. This dilution is equivalent to the standard accepted dilution of a needle content with 1 mL of 0.9% saline. Following dilution, the sample was sent to the Endocrine lab of SUMC where Tg concentration was evaluated by the commercial immunoassay, which is a solid phase two-site sandwich immunoassay, enzyme-labeled, chemiluminescent immunometric assay (Immulite 2000, System Analyzers, SIEMENS). POC-Tg kit performance was assessed retrospectively against the formal ELISA-based immunoassay results.

**Figure 3.**
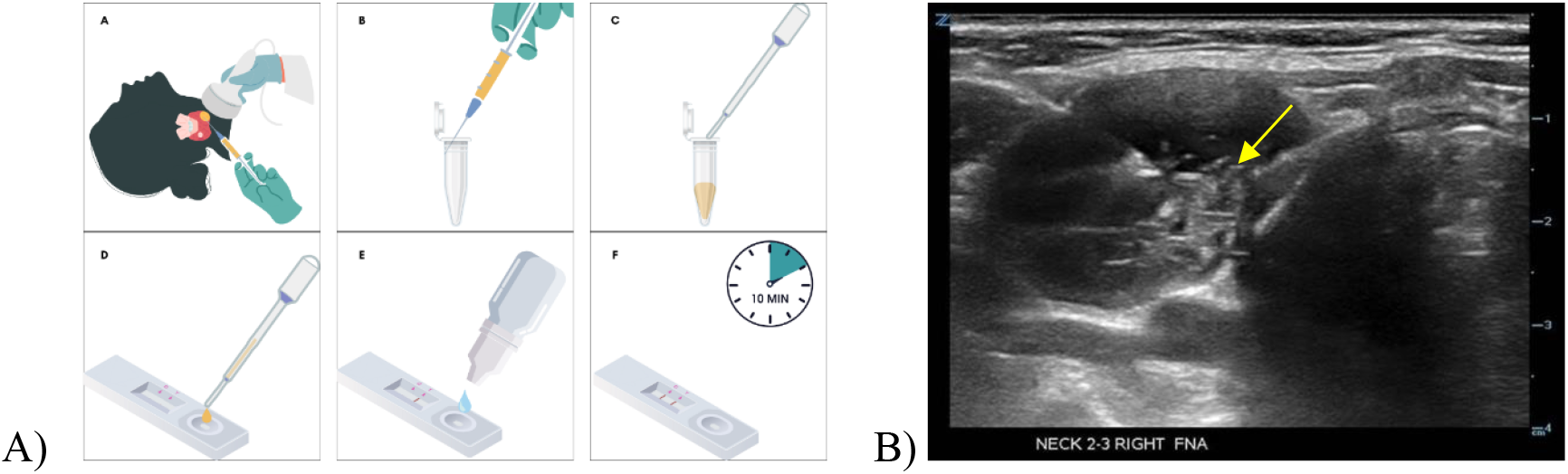
**A)** Schematic representation of the POC-Tg application procedure; Obtaining the sample during US-guided FNA procedure; Following dilution of the needle with 100 µl 0.9% w/w saline solution; The sample is transferred to an Eppendorf tube; 40µl is dispensed to the sample well of the test’s cassette; Two drops from the buffer dropper bottle are added to the well; Results are recorded and documented after 10 minutes. Image created with BioRender. **B)** FNA test of a suspicious lymph node in the right neck. In the depicted image, the needle can be seen puncturing through the lymph node, as highlighted by the yellow arrow.

**Figure 4.**
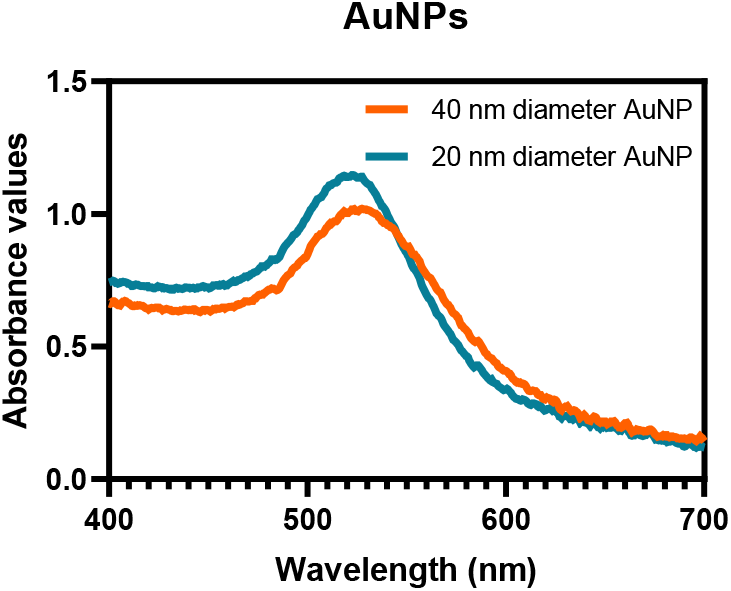
UV-Vis spectral analysis of colloidal gold confirming the presence of 20 nm and 40 nm diameter AuNPs.

As illustrated in Figure 3A, Once the FNA-Tg was collected and saline was added, the physiological fluid containing the putative target analyte was dispensed into the sample well, followed by the dispensing of a wash reagent buffer. The solution then reaches from the sample pad through the conjugate pad to the NC membrane, containing 2 lines of dispensed immunoglobulins. A reddish AuNP wet front seen as a wet stain moves towards the far end of the membrane. If the target analyte is present, then a visible line will be created in the test line location exhibiting a positive result. In the second line, which serves as a control indicator of the test’s integrity, a robust signal will be visible closer to the absorbent pad. A visible test line appears between 1-10 minutes after adding the sample and buffer, and this depends on the Tg concentration in the sample. Only positive results that appear within 10 minutes or less are considered true positives.

## 3. Results and discussions

The POC-Tg development process involved preparing AuNPs, conjugating antibodies, testing different antibody sources, selecting suitable membranes for optimal flow, and dispensing and characterizing antibody deposition on the membrane to achieve the desired LOD, free from false positives or negatives. After selecting the optimal combination of materials, the kits were validated using spiked and clinical samples in comparison with the NFAWF-Tg test. The results from these tests were compared to those obtained from clinical laboratory standards, using samples from the same specimens.

### 3.1. AuNP synthesis

A peak around 1 OD at the wavelength of 524-526 nm was observed, signifying the presence of 40 nm particles, which is the preferred size chosen for conjugation. An OD reading at 700nm was found to be 0.04, indicating absence of impurities. The solution was reevaluated via spectral analysis to confirm the absence of any undesirable changes.

### 3.2. Membrane selection

To determine the most suitable membrane for the device, three membrane combinations were assembled as described in the methodology. Membrane SN12 exhibited superior performance, with a stronger test line compared to both FF120 and CN150, as illustrated in Figure 5. The SN12 membrane consistently produced a clearer test line than the alternatives.

**Figure 5.**
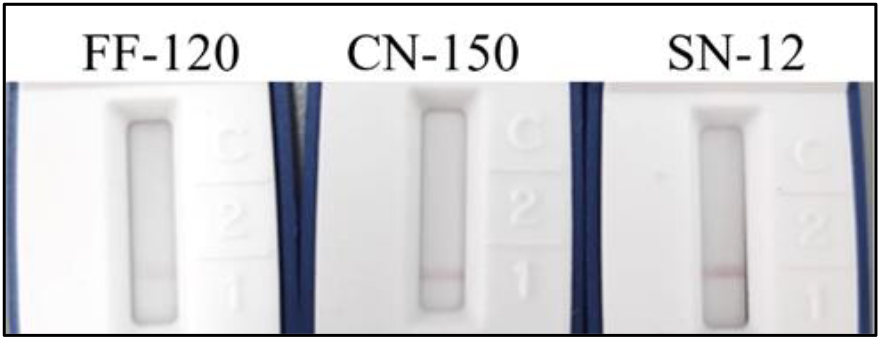
Evaluation of NC membranes (from left to right: FF120, CN-150, and SN12) through an elimination process. Three different membrane types were tested using a 5000 ng/ml Tg solution in PBS, with all other conditions kept consistent as outlined in the methods section.

### 3.3. Conjugation parameters

The final antibody concentration established by the cut-off evaluation was 3µg/ml/1 OD at pH 7. After synthesizing two different AuNP sizes and conjugating them with antibodies, only antibodies #395 and #392 demonstrated post-conjugation stability. These two gold-conjugated antibodies were then compared to identify the most suitable optimal choice for gold conjugate selection. Stability tests were conducted on multiple AuNP-conjugated antibodies (#392, #395, and a combination of #392 and #395), each tested at 1 OD on the selected SN12 membrane. Following testing, the strips were removed from the cassette for direct comparison. Phosphate-buffered saline (PBS) served as the negative control, while a 5000 ng/ml Tg solution in PBS was used as the positive control. Antibody #395 was identified as the optimal candidate for the conjugate pad, as shown in Figure 6.

**Figure 6.**
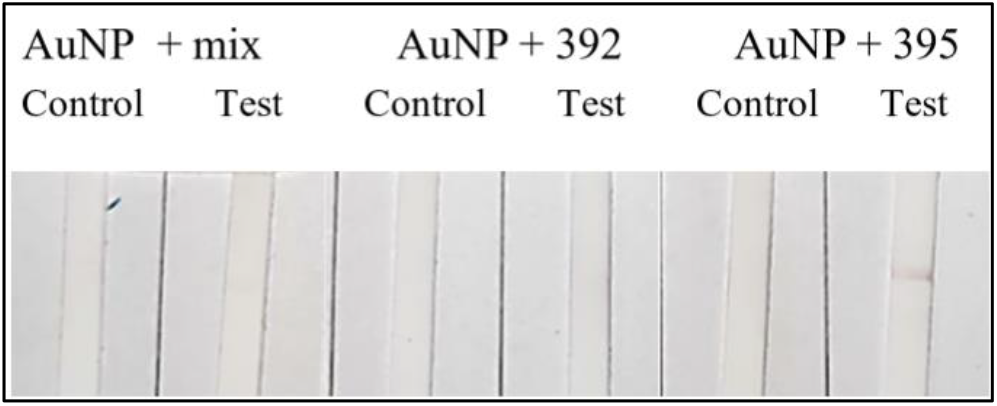
Comparison of three conjugate options for the device, with all other conditions consistent with those specified in the methods section. From left to right: AuNP+395&392 (mix) with PBS and Tg, AuNP+392 with PBS and Tg, and AuNP+395 with PBS and Tg.

**Figure 7.**
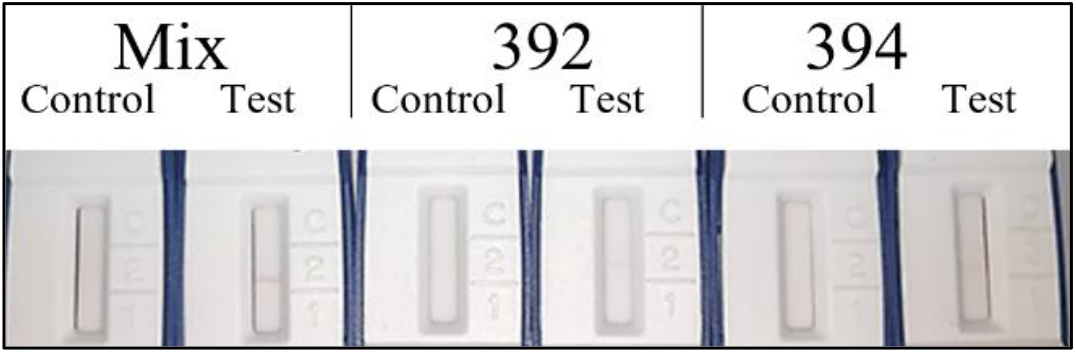
Comparison of three antibody combinations for the device, with all other conditions kept consistent as described in the methods section. PBS was used as the control, while a 5000 ng/ml Tg solution in PBS served as the positive test. From left to right: 392+393+394 (Mix) with PBS and Tg, 392 with PBS and Tg, and 394 with PBS and Tg. The mixed antibodies immobilized on the membrane produced the strongest signal, with a clearly visible line in the positive test. Both antibody 392 and antibody 394 exhibited weaker signals.

Regarding the conjugate pad performance, there was no significant difference between the 8980 and 6614 conjugation filter pads, as shown in Figure 8. Given that both demonstrated comparable performance, the 8980 pad was selected for the next optimization step. Following the optimizations, the optimal combination for the device comprises the combination of conjugation antibody: #395 at 1.0 OD, conjugation pad filter: 8980 and dispensed antibodies: #392+393+394 (Mix)

**Figure 8.**
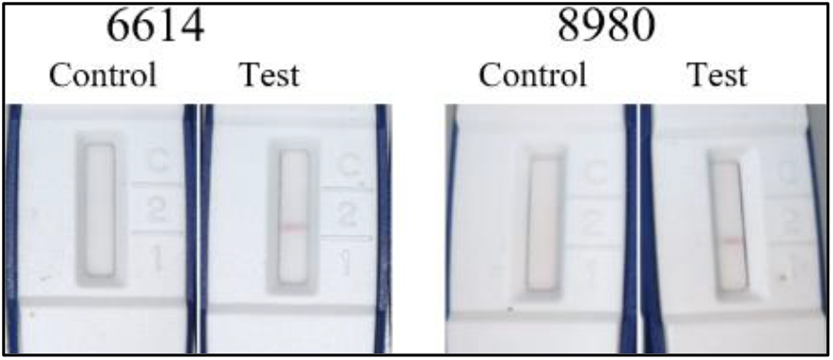
The performance of two conjugate filter pads, 6614 (left) and 8980 (right), was compared using PBS as the negative control, while 5000 ng/mL Tg in PBS served as the positive test.

### 3.4. Establishing the LOD

After optimizing the assay components, tests were conducted to establish the LOD using known Tg concentrations in human serum (Figure 9). Based on these results, only the samples that were tested directly were considered valid and added the test line strength vs Tg concentration analysis together with the standard clinical samples. The combined data was exclusively employed for assessing the correlation between the test line and Tg concentration in addition to the 30 clinical samples in table 1. Five replicates were tested at each Tg concentration—200 ng/mL, 100 ng/mL, 50 ng/mL, and 0 ng/mL (control)—in human serum to determine the LOD. The LOD was defined as the lowest concentration at which all replicate tests yielded a positive result for Tg, with any visible line (regardless of intensity) considered a positive indication.

**Table 1.** A comparison between the performance of the POC-Tg (presented as positive [P] or negative [N] answer) and the formal immunoassay. Samples were obtained during FNA procedure from a cervical LN suspicious as DTC metastases.

| Patient # | Sample name | POC-Tg P/N | the formal immunoassay | Line score |
| --- | --- | --- | --- | --- |
| 1 | 46 | P | > 30000 | 2 |
| 2 | 47 | P | > 30000 | 2 |
| 3 | 48 | P | 3604 | 0.5 |
| 4 | 7925 | N | Undetectable | 0 |
| 5 | 53 | N | Undetectable | 0 |
| 6 | 54 | N | Undetectable | 0 |
| 7 | 55 | P | 41.5 | 0.1 |
| 8 | 56 | P | > 30000 | 2 |
| 9 | 61 | N | Undetectable | 0 |
| 10 | 5413 | P | > 30000 | 1 |
| 11 | 64 | P | 26245 | 1.25 |
| 12 | 66 | P | > 30000 | 0.75 |
| 13 | 67 | P | > 30000 | 1.25 |
| 14 | 68 | N | Undetectable | 0 |
| 15 | 70 | P | > 30000 | 1 |
| 16 | 71 | N | 2.2 | 0 |
| 17 | 78 | N | Undetectable | 0 |
| 18 | 82 | N | Undetectable | 0 |
| 19 | 83 | P | 39.9 | 0.25 |
| 20 | 87 | P | > 30000 | 2 |
| 21 | 88 | N | Undetectable | 0 |
| 22 | 89 | N | Undetectable | 0 |
| 23 | 100 | N | Undetectable | 0 |
| 24 | 103 | N | Undetectable | 0 |
| 25 | 108 | P | 64.4 | 0.25 |
| 26 | 109 | N | 1.3 | 0 |
| 27 | 110 | N | Undetectable | 0 |
| 28 | 111 | P | > 30000 | 2 |
| 29 | 117 | P | 95.9 | 0.25 |
| 30 | 121 | P | > 30000 | 1.5 |

**Figure 9.**
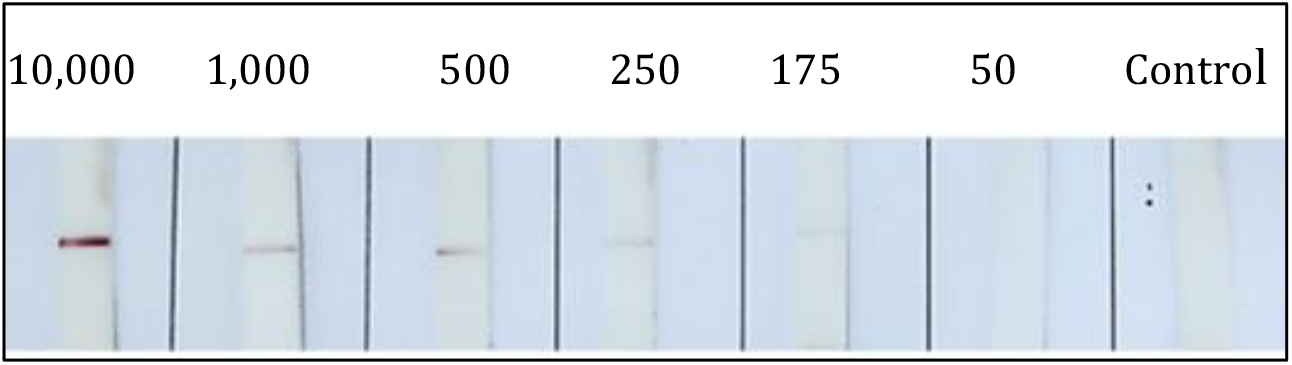
The spiking test (left to right) applied increasing Tg concentrations (ng/mL) of 10,000, 1,000, 500, 250, 175, 50, and 0 (negative control) to the sample wells.

**Figure 10.**
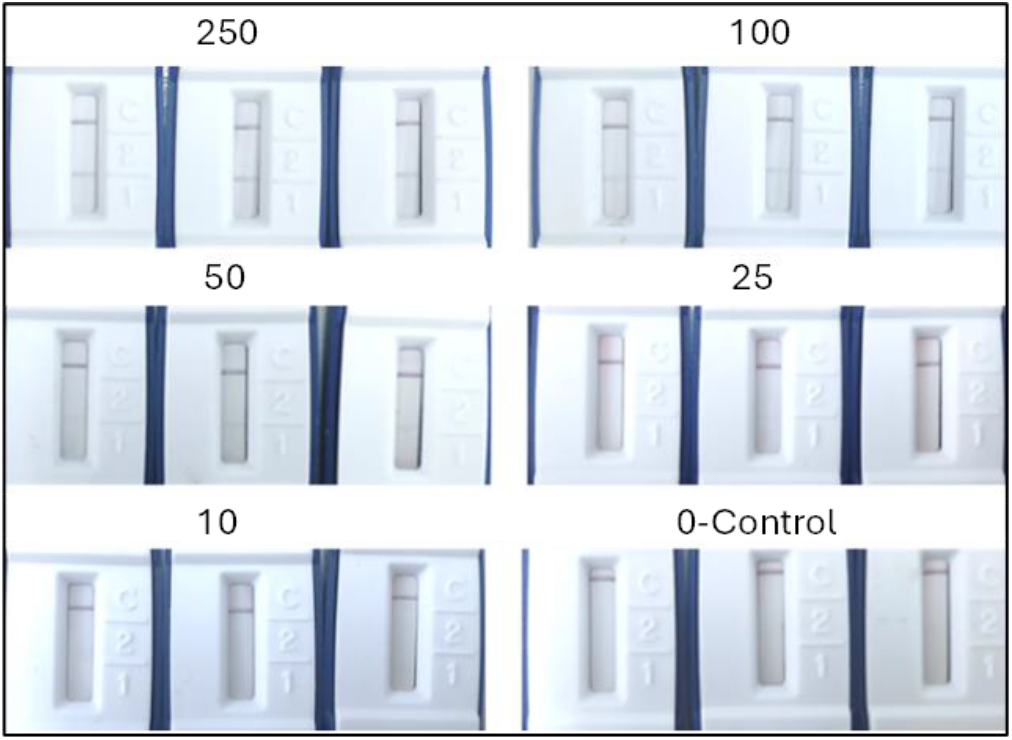
LOD test, using known concentration of Tg in serum. Top: triplicates of strips testing samples which contained 250 ng/ml (left), 3 samples with 100 ng/ml (right). Middle: triplicates of 50ng/ml (left) and triplicates of 25 ng/ml (right). Bottom: 10ng/ml (left) and 0ng/ml as control (right).

The conjugate OD was adjusted to 2.0 OD and the control line was strengthened by the following modifications. Firstly, we added 0.5 OD AuNP conjugated with Goat Anti rabbit IgG in the conjugate pad and then dispensed the control line on the NC membrane, which contained 0.5mg/ml of Rabbit anti human IgG. The following spiking tests were conducted using 3 repetitions for each concentration of 250, 100, 50, 25, 10 and 0 ng/ml of Tg in serum. Throughout all the tests with 250ng/ml and 100ng/ml there were visible test lines. However, in lower concentrations of 50 ng/ml, all samples showed a faint positive signal. In lower concentrations of 25ng/ml and 10ng/ml, no signal was observed.

### 3.5. Clinical validation

The LOD test indicates that the LOD is indeed 50ng/ml, which is the equivalent concentration to the desired 5ng/ml of the thirty samples obtained during FNA-Tg of a suspicious cervical LN were tested with both the POC-Tg and the standard immunoassay. The maximal concentration reported by the standard immunoassay is 30,000ng/ml, and concentration above this value were recorded as > 30,000 ng/mL. Fourteen samples were found to be negative and sixteen samples were positive, using our POC-Tg tests. In 12 of the 14 negative results both the POC-Tg test and Immulite 2000 reported undetectable Tg as well. Two samples showed in Immulite 2000 a low but detectable Tg: 1.33 and 2.2 ng/ml, both below the LOD for which the POC-Tg was set. All 16 positive samples by the POC-Tg gave a positive result by the formal immunoassay. Ten of them reached the maximum reading of 30,000 ng/ml, and 4 of them showed a reading of less than 100 ng/ml (41.5, 39.9, 64.4 and 95.9 ng/ml). Accordingly, the POC-Tg was judged to have specificity and sensitivity of 100% when compared with the formal immunoassay. Regarding the two samples that exhibited a low value beneath the predetermined cut-off threshold, a subsequent cytological analysis for the first sample reported metastatic PTC. The second was non-diagnostic. Finally, another ultrasound examination conducted by a radiologist characterized this LN as benign. In total, the diagnostic accuracy in terms of final diagnostic was 96.6%.

The strength/intensity of the test line was assessed utilizing the test line score color legend. Several clinical samples with different test line scores are shown in Figure 11.

**Figure 11.**
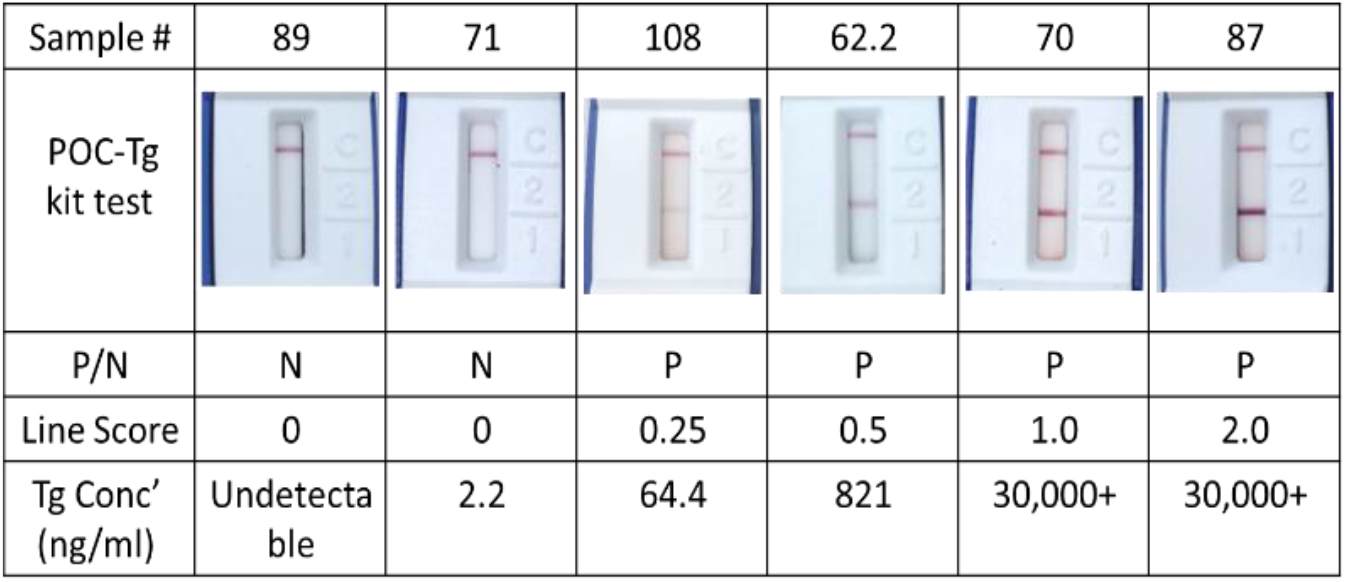
Examples of positive and negative clinical samples obtained after FNAWF-Tg with range of signals line score in the POC-Tg kit. From top to bottom: the sample number, POC-Tg test picture, P/N-positive or negative results according to POC-Tg, test line score of the test, and Tg concentration in ng/ml according to standard ELISA immunoassay.

**Figure 12.**
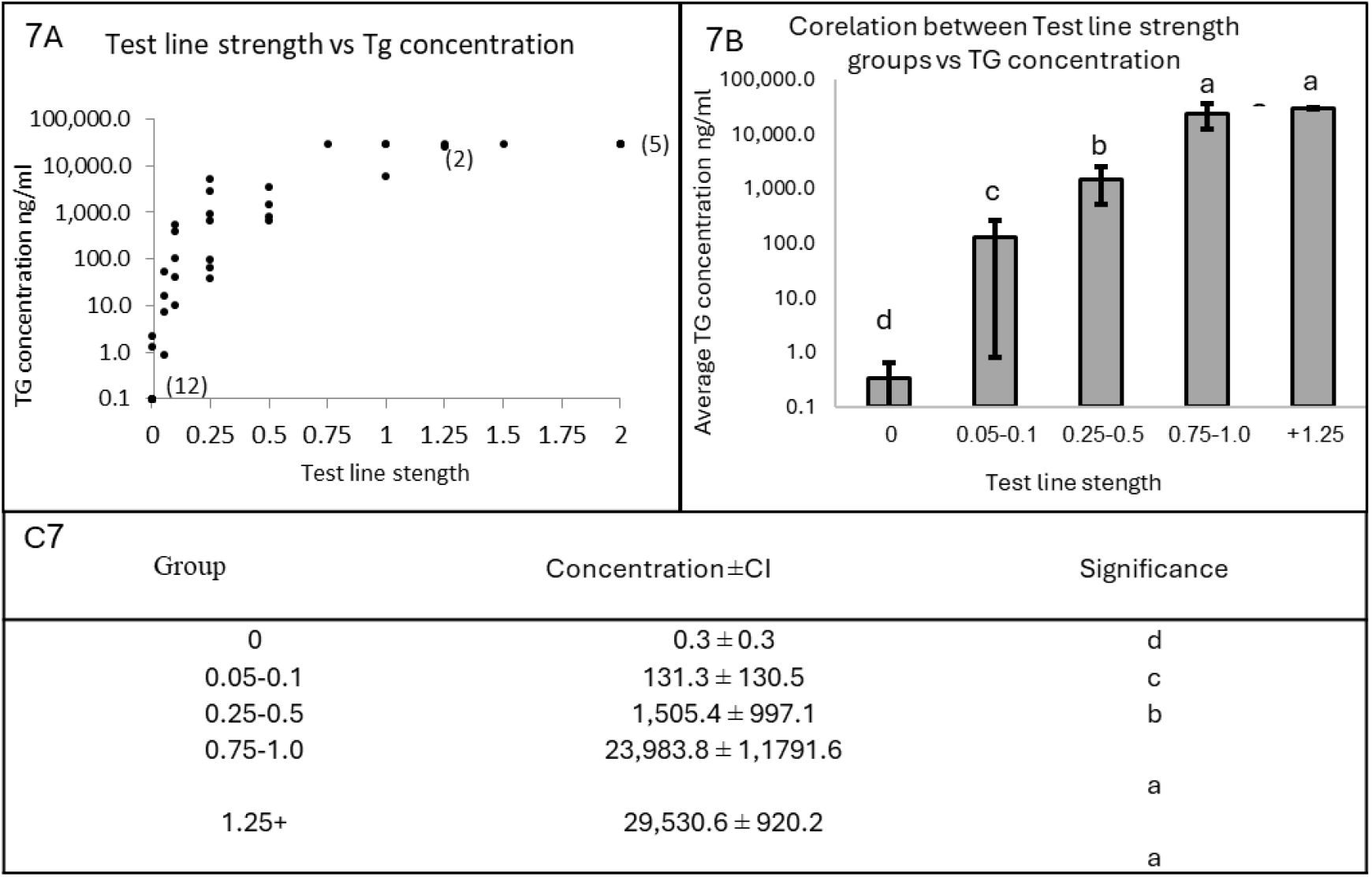
Relationship between the test line strength and Tg concentration. Graph 7A shows the relationship between test line strength and Tg concentration. In 9 out of 10 samples with Tg concentrations exceeding 30,000 ng/mL, the test line strength was 1.0 or higher. Graph 7B demonstrates the correlation between test line intensity and average Tg concentration, with samples grouped by test line score. The average Tg concentration is shown in logarithmic scale. Table 7C displays significant differences between groups of test line score.

The samples in Figure 11, were tested in different times and the pictures were taken 10 minutes after running the test. From left to right: sample 89 was found negative using our test and undetectable in the standard test. Sample 71 was found negative using our test and 2.2 ng/ml in the standard test-meaning a negative result. Sample 108 resulted positive (score of 0.25) using our test and 64.4 ng/ml in the standard test. Sample 62.2 resulted positive (score of 0.5) using our test and 821 ng/ml in the standard test. Sample 70 was found positive (score of 1.0) using our test and over 30,000 ng/ml in the standard test. Sample 87 results positive (score of 2.0) according to our test and over 30,000 ng/ml in the standard test.

To test the correlation between the strength of the test line displayed through LFIA POC-Tg and the concentration of Tg measured using Immulite 2000, the clinical samples (n=46) were divided into test line strength groups. The mean concentrations of Tg for each group were analyzed to ascertain significant differences between them. The 12 clinical samples with undetectable concentrations of Tg counted as 0.1 ng/ml while every concentration higher than 30,000 ng/ml was capped at 30,000 ng/ml. The average Tg concentration per group was measured with a confidence interval (CI) of 95%. The groups were tested with Student’s t-Test for significant differences amongst them, and every significantly different group was labelled with letter (p<0.05). The findings demonstrate a correlation between the strength of the test line and the corresponding TG concentration. There is a significant difference for every group except for 0.75-1.0 and 1.25+, which showed no significant difference. These results suggest that the POC-Tg exhibits semi-quantitative characteristics, allowing for a rough estimation of Tg concentration based on the strength of the test line.

## 4. Conclusions

This study demonstrates significant progress in developing a POC-Tg designed for the detection of DTC metastasis involving cervical LNs. The semi-quantitative POC-Tg was optimized to attain a LOD of 5ng/ml following dilution, designated as the threshold for a positive result. A cohort of 30 clinical samples was procured via FNA-Tg procedure from patients with suspected cervical LN. The POC-Tg exhibited 16 positive results and 14 negative results. The test showed 100% specificity (n=14) and sensitivity (n=16) when compared to the standard ELISA-based test, demonstrating robust performance with the tested clinical samples. All the negative results were either undetectable in the standard ELISA test or fell below the 5ng/ml limit. All the positive results obtained with the POC-Tg surpassed the established limit of 5ng/ml. The POC-Tg demonstrated 96.6% diagnostic accuracy for the final detection of cervical LN metastases of DTC origin. Results obtained in the clinic from suspected lymph nodes show that there is a correlation between the strength of the test line and the Tg concentration. Further research to lower the LOD of the POC-Tg (e.g., LOD equal to 1 ng/mL) may be of value where there is a gap between a highly suspicious LN and a negative result by the POC-Tg. We believe that broader testing and optimization can further lower the threshold of sensitivity required for any given necessity within the 1-10 ng/ml range. This test could easily be performed and used at the clinic during the FNA-Tg procedure, offering rapid results that could enhance clinical decision-making and ease the healthcare burden. To further validate our POC-Tg, a comprehensive multi-center clinical study is needed to test the kit with a broader number of patients and formal samples.

## Data Availability

All data produced in the present study are available upon reasonable request to the authors

## Future directions

The next stage of this study involves conducting an inter-institutional study, collaborating with a network of endocrine physicians attending to the same type of patients. This approach will allow for the validation of the POC-Tg by independent clinicians throughout the multi-center study, ensuring an assessment of its efficacy beyond the developers of the test.

## Author Contributions

Conceptualization, U.Y., M.F., and R.S.M.; investigation and analysis, S. A., K.K., E.B., M.E., T.A., and U.Y.; writing, S.A., U.Y., K.K., and R.S.M.; review and editing, S.A, K.K., M.F., E.B., M.E., T.A., U.Y., and R.S.M. All authors have read and agreed to the published version of the manuscript.

## Patent

This research was patented through BG-Negev: “Devices and methods for diagnosing thyroid medical conditions.” U.S. Provisional Patent Application Publication No. 63/138,481.

## Acknowledgments

We extend our gratitude to Dr. Amos Sommer and to Mr. Ron Ramon, whose expertise and willingness to share their knowledge enriched our research process. Additionally, we extend our deepest thanks to Mrs. Maayan Angel. Her encouragement and patience have been irreplaceable to the first co-author Mr. Sagi Angel, providing the necessary environment for focus and dedication. We thank the Israel Cancer Association for the generous grant that supported this research (grant number 20230021).

